# Predictive factors of response to 3^rd^ dose of COVID-19 mRNA vaccine in kidney transplant recipients

**DOI:** 10.1101/2021.08.23.21262293

**Authors:** Xavier Charmetant, Maxime Espi, Thomas Barba, Anne Ovize, Emmanuel Morelon, Olivier Thaunat

**Author notes:** **Correspondence to:** Olivier Thaunat MD, PhD, Service de Transplantation, Néphrologie et Immunologie Clinique, Hôpital Edouard Herriot, 5 Place d’Arsonval, 69003 Lyon, France. equal contributions (Xavier Charmetant, Maxime Espi).

## Abstract

Only a minority of kidney transplant recipients (KTRs) develop protective neutralizing titers of anti-receptor binding domain of spike protein (RBD) IgG after two doses of mRNA COVID-19 vaccine. Administration of a third dose of mRNA vaccine to KTRs with sub-optimal response increase anti-RBD IgG titers but with high inter-individual variability. Patients with the higher response rate to the third dose of vaccine can be identified by the presence of low anti-RBD IgG titers and spike-specific CD4+ T cells in their circulation 14 days after the second dose.

## Introduction

Kidney transplant recipients (KTRs) carry a very high risk of death due to COVID-19 in case of infection by SARS-Cov-2 ^1–5^. This vulnerable population has therefore been prioritized for vaccination. However, after the “standard” 2 doses of mRNA vaccine only few KTRs develop appropriate levels of anti-receptor binding domain of spike protein (RBD) IgG ^6–8^, which are responsible for the neutralization of the virus and the protection against COVID-19 ^9^. Accordingly, we and others have reported cases of vaccinated KTRs with (sometime severe forms of) COVID-19 ^10,11^.

Preliminary reports suggest that a third dose (D3) of vaccine may improve the humoral response in some patients ^12,13^. In this prospective observational study, we aimed at describing the serological response of KTRs to D3 of mRNA vaccine and identifying the variables associated with response to D3.

## Short methods

### Study population

The study protocol was approved by Institutional Review Board (approval number: 2020-A02918-31, Comité de Protection des Personnes Sud-Est I). Vaccination with 2 doses of BNT162b2 mRNA vaccine (Pfizer-BioNtech) was offered to KTRs from Lyon University Hospital. According to the French health authority, a third vaccine injection was offered to all patients whose IgG titers were below 142 BAU/mL, the threshold correlating with positivity of functional neutralizing assay ^14^.

### Anti-SARS-Cov2 S-RBD humoral response assessment

The IgG antibodies directed against the Receptor Binding Domain (RBD) of the spike glycoprotein of the SARS-Cov2 were detected by a chemiluminescence technique, using the Maglumi® SARS-CoV-2 S-RBD IgG test (Snibe Diagnostic, Shenzen, China) on a Maglumi 2000® analyser (Snibe Diagnostic), according to the manufacturer’s instructions. This test displays clinical sensitivity and specificity of 100% and 99.6%, respectively. As recommended by the WHO, the obtained titer was then expressed as binding arbitrary units/mL (BAU/mL); correction factor for Maglumi®: 4.33.

### Anti-SARS-Cov2 Spike cellular response assessment

Spike specific CD4+ T cells response was quantified in the circulation of the KTRs using the QuantiFERON® SARS-CoV2 test (Qiagen, Netherlands), a commercially available Interferon Gamma Releasing Assay (IGRA), according to the manufacturer’s instructions.

### Statistical analysis

All the analyses were carried out using R software version 4.0.4 (R Foundation for Statistical Computing, Vienna, Austria, 2021, https://www.R-project.org) and or GraphPad Prism v8.0 (San Diego, California USA).

## Results

Anti-RBD IgG and the interferon-γ secreted by circulating spike-specific CD4+ T cells were monitored 14 days after the second dose (D2) of BNT162b2 vaccine (Pfizer-BioNtech) in 93 consecutive KTRs from Lyon University Hospital, 5 of whom had previous history of COVID-19 (black diamonds; **Figure 1A-C**).

**Figure.**
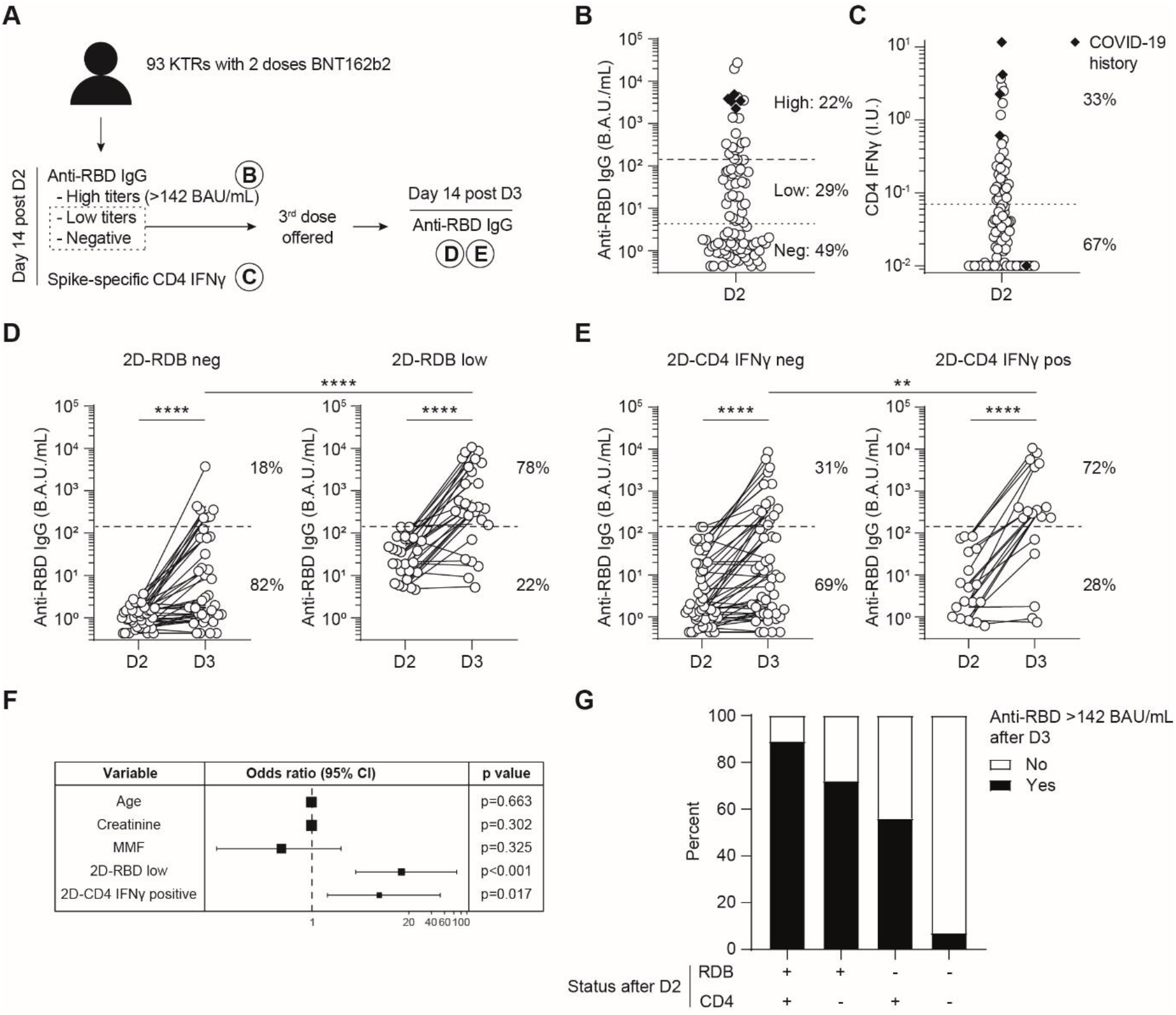
**A**. Experimental design of the clinical observational study. **B-C**. Antibodies directed against the receptor binding domain (RBD) of the spike protein of SARS-CoV-2 (anti-RBD IgG) and the interferon-γ secreted by circulating spike-specific CD4+ T cells (CD4 IFNγ) were monitored 14 days after the second dose. Black diamonds represent patients with a previous history of COVID-19. **B**. Titers of anti-RBD IgG. Dotted line indicates the threshold of positivity of the assay, dashed line indicates the threshold of 142 BAU/mL, considered as protective against COVID-19. **C**. Interferon-γ secreted by spike-specific CD4+ T cells. **D-E**. Titers of anti-RBD IgG after D3 according to the absence (left panel) or presence (right panel) of low titers of anti-RBD IgG (2D-RBD) (**D**) or circulating spike specific CD4+ T (CD4 IFNγ) cells (**E**) after D2. **F**. A multivariate analysis was conducted to identify the variables independently associated with a titer of anti-RBD IgG > 142 BAU/mL after D3. Forest plot shows the odd ratio and the 95% confidence interval for the variables with p<0.10 in the univariate analysis. **G**. The probability to develop a protective titer of anti-RBD IgG after D3 is shown according to the result of the two predictive biomarkers after D2, i.e. low titers of anti-RBD IgG (RBD) or presence of circulating spike specific CD4+ T cells (CD4). **Abbreviations are:** KTRs: kidney transplant recipients; RBD: receptor binding domain; D2: second dose; D3: third dose; BAU: binding antibody unit. Mann-Whitney test for unpaired data and Wilcoxon test for paired data. **, p<0.01; ****, p<0.0001.

Following French health authorities recommendation, a third dose (D3) of vaccine was injected to 66 patients of the initial cohort, (dashed line in Figures 1B-E). Overall, the clinical tolerance of the D3 was good. The main side-effect was pain at the site of injection, which occurred with the same incidence after each of the three doses (≈50% of patients). Five patients had fever <39°C for maximum two days after D3.

Forty-two percent (28/66) of KTRs that received D3 reached 142 BAU/mL of anti-RBD IgG (responders). In univariate analysis (**Table 1**), responders to D3 were younger than non-responders (43.2+/-13.1 vs 52.1+/-12.5 years; p=0.011), had lower baseline creatinine level (114+/-33 vs 143+/-50 µmol/L; p=0.015), and were less frequently exposed to mycophenolate mofetil (19/28 vs 35/38, p=0.021). In addition, presence of low titers (i.e. above threshold of positivity but below 142 BAU/mL) of anti-RBD IgG (**Figure 1D**), and circulating spike-specific CD4+ T cells (**Table 1, Figure 1E**) after D2, were both associated with a better serological response to D3. In multivariate analysis, these last two biomarkers were the only remaining independent predictive variables allowing identifying KTRs responders to D3 (OR 15.9, IC_95%_ [3.87−87.46], p<0.001, for low anti-RBD IgG titers; and 8.03, IC_95%_ [1.62−52.59], p=0.017 for positive CD4+ T cells IGRA; **Figure 1F**). Furthermore, combining these two information, we observed that the probability to respond to D3 in KTRs was the highest in patients positive for both tests (89%, **Figure 1G**). The response rate decreased to 72% in KTRs with only low anti-RBD IgG after D2 and 56% in those with only a positive CD4+ T cells IGRA. Finally, only 7% KTRs in whom both tests were negative after D2 did develop an optimal serological response after D3.

**Table.** Characteristics of patients from Lyon University Hospital cohort.

| n (%) or mean +/- SD | Initial cohort<br>(2 doses)<br>n = 93 | KTRs who received D3 (n=66) |  | p* |
| --- | --- | --- | --- | --- |
|  |  | Non-responders to<br>D3<br>n = 38 | Responders to<br>D3<br>n = 28 |  |
| **Age (y)** | 56.3 ± 12.3 | 60.4 ± 11.0 | 53.8 ± 13.3 | **0.032** |
| **Male** | 50 (54) | 20 (54) | 17 (46) | 0.514 |
| **Time from transplantation (y)** | 10.4 ± 8.8 | 8.3 ± 7.0 | 10.6 ± 9.5 | 0.255 |
| **Immunosuppressive drugs** |  |  |  |  |
| CNI | 85 (91) | 36 (95) | 25 (89) | 0.643 |
| MMF/MPA | 71 (76) | 35 (92) | 19 (68) | **0.018** |
| Steroids | 80 (86) | 33 (87) | 24 (86) | 0.999 |
| imTOR | 8 (9) | 1 (3) | 4 (14) | 0.154 |
| Belatacept | 1 (1) | 1 (3) | 0 (0) | 0.999 |
| **Biological data** |  |  |  |  |
| Lymphocytes (G/L) | 1.6 ± 0.8 | 1.4 ± 0.7 | 1.7 ± 0.7 | 0.238 |
| Monocytes (G/L) | 0.7 ± 0.2 | 0.7 ± 0.2 | 0.6 ± 0.1 | 0.928 |
| Creatinine (μmol/L) | 126 ± 46 | 143 ± 50 | 114 ± 33 | **0.015** |
| **COVID-19 history** | 5 (5) | 0 (0) | 0 (0) | - |

\*: qualitative variables were compared using a two-sided Chi-square test or a two-sided Fisher's exact test when the conditions for a Chi-square were not fulfilled, quantitative variables were compared using a t-test.
**Abbreviations are:** y, years; CNI, calcineurin inhibitor; MMF/MPA, mycophenolate mofetil/mycophenolic acid; imTOR, inhibitor of the mechanistic target of rapamycin.

## Discussion

A 3^rd^ dose of BNT162b2 significantly increases the titers of anti-RBD IgG in KTRs that did not develop a protective antibody response after the “standard” 2 doses scheme of vaccination. In line with previous studies ^12,13,15^, although well tolerated, response of KTRs to D3 appeared highly heterogeneous with only 42% (28/66) of patients considered as protected after D3. Identification of patients that would benefit from D3 is therefore an important unmet medical need in order to optimize the protection of this vulnerable population while avoiding wasting precious vaccine doses. This study provides preliminary indications that the discrimination between responders and non-responders to D3 could be achieved by measuring anti-RBD IgG and spike-specific CD4+ T cells in the circulation of KTRs 14 days after D2. Based on these results, it seems logical to not propose D3 to KTRs who have developed neither anti-RBD IgG nor spike-specific CD4+ T cells after D2. The latter, might instead rather benefit from passive immunization with anti-SARS-Cov-2 monoclonal antibodies, as recently suggested in another vulnerable population of residents and staff in assisted living facilities ^16^.

## Data Availability

NA

## Acknowledgements

The authors are indebted to the members of the GRoupe de REcherche Clinique (GREC: Céline Dagot, Farah Pauwels, Fatiha M’Raiagh and Daniel Sperandio) for excellent technical assistance during the collection of the samples, and to the clinicians (Fanny Buron, Charlène Lévi, Alice Koenig, Marion Delafosse and Louis Manière) who recruited the patients. OT is thankful to Christine Bouz, Laurence Pellegrina, Emmanuelle Cart-Tanneur, Lise Siard, Claudine Lecuelle, and Philippe Favre from Eurofins Biomnis for their help during the conduction of the study.

XC is supported by a funding from the Société Française de Transplantation. ME is supported by the Hospices civils de Lyon and currently holds a “poste accueil” position funded by INSERM. OT is supported by the Etablissement Français du Sang and the Fondation pour la Recherche Médicale (PME20180639518).

## Supplementary materials

### Detailed methods

#### Anti-SARS-Cov2 S-RBD humoral response assessment

Ten microliters of serum were incubated in the appropriate buffer with magnetic microbeads covered with S-RBD recombinant antigen, in order to form immune complexes. After precipitation in a magnetic field and washing, ABEI (N-(4-Aminobutyl)-N-ethylisoluminol)-stained anti-human IgG antibodies were added to the samples. After a second magnetic separation and washing, the appropriate reagents were added to initiate a chemiluminescence reaction. When necessary, sera were diluted sequentially up to 1:1000.

#### Anti-SARS-Cov2 Spike cellular response assessment

One milliliter blood was distributed in each tube of the assay: (i) uncoated tube: negative control/background noise, (ii) tube coated with mitogen: positive control, and (iii) tube coated with HLA-II restricted 13-mers peptides derived from the entire SARS-CoV2 Spike glycoprotein used to stimulate CD4+ T cells. After 20 hours of culture at 37°C, tubes were centrifugated 15 minutes at 2500g, and stored at 4°C before INFγ quantification in the supernatant by ELISA.

The CD4+ T cell assay value was the difference between tube (iii) and the negative control (i).

#### Statistical analysis

Categorical variables were expressed as percentages and compared with a two-sided chi-square test or a two-sided Fisher’s exact test when the conditions for a chi-square were not fulfilled. Continuous variables were expressed as mean ± SD and compared using t-tests or as median and compared using Mann Whitney test for variables with non-normal distribution. Wilcoxon test was used for paired data.

Logistic regression models were used in both univariate and multivariate analyses. All the explanatory variables significantly associated with outcomes in univariate analyses (p-value < 0.10) were included in multivariate models. Stepwise regression analyses with bidirectional elimination were then performed, using Aikake Information Criterion to select the most fitting final multivariate models.

